# Camp Dream. Speak. Live.: A Virtual Adaptation

**DOI:** 10.1101/2024.08.22.24311294

**Authors:** Courtney T. Byrd, Geoffrey A. Coalson, Danielle Werle, Robyn Croft, Katie L. Winters, Megan M. Young

**Author notes:** Address for correspondence: Geoffrey Coalson, Speech, Language, and Hearing Sciences, The University of Texas at Austin, CMB 3.102, Austin, TX 78759, USA.

## Abstract

**Purpose:** The purpose of this study was to determine the efficacy of a virtual adaptation of the administration of Camp Dream. Speak. Live., an intensive, non-ableist manualized treatment program for children who stutter, with no indirect or direct fluency goals, in reducing the adverse impact of stuttering and increasing communication competence.

**Methods:** Sixty-one children who stutter participated in Virtual Camp Dream. Speak. Live. Pre- and post-treatment measures were identical to previous in-person administrations: (1) self- and caregiver-report of cognitive and affective impact of stuttering (*Communication Attitude Test for Children who Stutter* [*KiddyCAT/CAT*], *Overall Assessment of Speaker’s Experience of Stuttering* [*OASES*], *PROMIS Pediatric Peer Relationship,* and *PROMIS Parent Proxy Relationships*), and (2) unfamiliar clinician ratings of communication competence of impromptu presentations.

**Results:** Significant post-treatment gains were reported for the *CAT*, *OASES*, and *PROMIS Peer Relationships Parent Proxy*. Significant gains in post-treatment communication competence were observed. Pre-treatment stuttering frequency did not significantly predict changes in communication competence.

**Conclusion:** Findings from Virtual Camp Dream. Speak. Live. demonstrate that the administration of the adapted telepractice format of this manualized program yields comparable findings as when administered in-person, suggesting promising implications for use in locations for which in-person provision and/or access is not feasible.

## 1.0 Introduction

The COVID-19 pandemic permanently changed the landscape of allied healthcare professions. Telepractice has become a more acceptable alternative to in-person treatment within speech-language pathology (American Speech-Language-Hearing Association [ASHA], n.d.). A number of benefits of telepractice have been documented (e.g., scheduling flexibility, elimination of geographical barriers, time and cost benefits, caregiver involvement, natural environment), as well as potential drawbacks (e.g., limited evidence base, professional training and licensure, equipment and connectivity expense, technological barriers between generations and socioeconomic strata; Raatz et al., 2021; Theodoros, 2011). Given the ever-increasing use of telepractice in clinical service provision, there is a critical need to investigate the effectiveness of adapting manualized, in-person treatment programs to virtual formats and to determine whether similar outcomes are achieved.

Camp Dream. Speak. Live. is a one-week intensive treatment program offered to children and adolescents who stutter in multiple countries (e.g., Belgium, Ireland, Israel, Malta, Pakistan, Poland, The Netherlands, United States), during which the participants engage in communication, advocacy, resiliency, and educational activities (see Blank Center CARE^™^ Model). Previous clinical research has indicated post-intervention gains across cognitive and affective components of stuttering and communication effectiveness (Byrd et al., 2016a, 2016b, 2018, 2021, *N* = 83 total participants). To date, empirical studies investigating the outcomes of Camp Dream. Speak. Live. have only examined the outcomes of in-person participation. The purpose of the present study is to assess whether previously documented post-intervention findings are observed in a virtual adaptation of Camp Dream. Speak. Live.

### 1.1 History of telepractice for children and adults who stutter

Since 2005, ASHA has supported the use of telepractice for speech-language pathology services, with the expectation that it has the potential to provide clinical outcomes comparable to in-person treatment (ASHA, 2005; Brown, 2011). Prior to the COVID-19 pandemic, the use of telepractice for children who stutter produced a limited, but promising number of small-scale studies supporting its clinical efficacy in one-on-one and in group delivery.

#### 1.1.1 One-on-one telepractice sessions

The majority of the studies that have investigated one-on-one telepractice for individuals who stutter have provided data from small cohorts of variable age ranges (Carey et al., 2012 [*n* = 3, 13 to 16 years of age]; Kully, 2000 [*n* = 1, 38 years of age]; O’Brian et al., 2014 [*n* = 3, 3 to 4 years of age]; Sicotte et al., 2003 [*n* = 6, 4 to 19 years of age]; Valentine, 2014 [*n* = 2, 11 years of age]). A single-arm clinical trial by Carey et al. (2014, *n* = 14, ages 12 to 17) examined the outcomes of the Camperdown Program via synchronized, one-on-one webcam sessions which provided modeled examples of speech fluency techniques for adolescents who stutter (i.e., prolonged speech), along with post-session email correspondence (range of 7.3 to 26.3 hours of treatment per child). A significant reduction in percentage of stuttered syllables (%SS), self-rated stuttering severity, situational avoidance, and adverse impact of stuttering was reported 12-months post-treatment, although notable variability was present across all outcome measures.

A parallel-arm clinical trial by Bridgman et al. (2016) investigated the clinical efficacy of the Lidcombe Program for preschool-aged children (3 to 5 years of age) wherein caregiver consultations were randomly assigned to be administered in-person (*n* = 24) or via webcam (*n* = 25). Similar to Carey et al. (2014), significant changes in %SS and caregiver-reported stuttered speech were detected for both in-person and telepractice conditions from pre-treatment to 18-months post-treatment. No cognitive or affective measures following telepractice sessions were obtained for these preschool-aged participants.

Recently, Eslami Jahromi et al. (2022) administered the Camperdown Program to a cohort of adolescents and adults who stutter (*n* = 30, 14 to 49 years of age) via one-on-one, synchronous virtual sessions. Eslami Jahromi and colleagues also reported significant post-treatment reductions in stuttered speech as measured by %SS and the *Stuttering Severity Instrument*-*Fourth Edition* (*SSI-4*; Riley, 2009), but, similar to Bridgman et al. (2016), no cognitive or affective measures were examined pre- or post-treatment.

Taken together, from the limited number of studies investigating one-on-one telepractice for individuals who stutter, virtual administration appears to yield comparable findings as in-person administration. That being said, these one-on-one telepractice sessions and post-treatment outcomes predominantly focused on increasing fluency and/or modifying moments of stuttered speech. Further, one-one-one telepractice sessions may not provide the same benefits as group telepractice sessions (e.g., Hearne et al., 2008).

#### 1.1.2 Group telepractice sessions

Few studies have examined the efficacy of group telepractice for persons who stutter, and when examined, group sessions have either accompanied or followed one-on-one treatment sessions. For example, Tomaiuoli et al. (2021) reported clinical outcomes for children who stutter (*n* = 11, *M* age = 9 years old) following group telepractice sessions targeting fluency shaping and stuttering modification, with the purpose of comparing these findings to their prior in person administration with an age-matched sample. In this study, children completed biweekly one-on-one telepractice sessions (fluency shaping, stuttering modification), followed by weekly group treatment sessions. Data indicated an overall reduction in stuttering severity, as measured by the *SSI-4*, and adverse impact of stuttering, as measured by the *Overall Assessment of Speaker’s Experiences of Stuttering* (*OASES;* Yaruss & Quesal, 2016). Tomaiuoli and colleagues reported that these telehealth findings were comparable to their prior in-person administration.

Cangi and Toğram (2020) examined virtual group treatment for adults who stutter (*n* = 20; *M* age = 28 years). Treatment consisted of the Therapy Stage, which included nine individual sessions and three group sessions focusing on fluency shaping and stuttering modification, followed by the Maintenance Stage, which included six group sessions to encourage participants to continue using their strategies for stuttering. Adult participants completed treatment and maintenance either entirely in-person (*n* = 10) or via telepractice (*n* = 10). Cangi and Toğram reported significant changes in stuttering (as measured by %SS and *SSI-4*) and affective measures (as measured by the *OASES* and the *St. Louis Inventory of Life Perspectives and Stuttering* [St. Louis, 2001]). Similar to Tomaiuoli et al. (2021), no difference was detected between in-person and telepractice formats four months post-treatment, suggesting comparable clinical outcomes across administration formats.

Although the limited number of studies suggest that outcomes for group telepractice for individuals who stutter are similar to in-person formats, further examination with larger cohorts is warranted. Given the greater challenges with facilitating group exchanges within a virtual setting, additional research is particularly needed for virtual adaptations of group treatment formats. Moreover, the majority of telepractice studies with people who stutter have examined treatment approaches wherein increasing fluency and/or stuttering more easily is a direct or indirect clinical outcome. Virtual administration of non-ableist clinical programs that do not view stuttering as a condition that needs to be fixed (see Byrd, 2021; Constantino et al., 2022; Venkatagiri, 2009; Watermeyer & Kathard, 2016) have yet to be examined.

### 1.2 In-person group treatment for children who stutter

Some clinical programs for children who stutter provide support/self-help within an overnight, recreational summer camp environment that do not include formal treatment goals for all participants (e.g., Camp SAY, see Herring et al., 2021). Others are a hybrid format in that they offer a recreational camp experience while also incorporating treatment goals for participants, including explicit fluency-focused and/or stuttering modification goals (see Byrd et al. [2016a] for review). For example, during Camp Shout Out, on “Attentive Tuesday - …[campers] can modify the speaking process, utilizing an easier relaxed speech, phrasing and pausing, …or modifications of the stuttering moment” (Byrd et al., 2016a, p. 60), and at Camp TALKS, on “Day 2 - …campers challeng[e] themselves to say the words they want to say or use speech tools [such as] pull-outs, soft starts, easy stutters, etc.” (p. 64).

Camp Dream. Speak. Live. (e.g., Byrd et al., 2016a, 2018, 2021) is not a traditional or quasi-recreational summer camp; the word “camp” is used to make it more inviting for children. Camp Dream. Speak. Live. has a manualized protocol that has been administered in-person by other clinicians and researchers across several locations nationally and internationally. Scripted daily activities directly target the four aspects of the Blank Center CARE^™^ Model: Communication, Advocacy, Resiliency, and Education (see Byrd et al. [2016a, 2016b] for a description of Camp Dream. Speak. Live., see https://blankcenterforstuttering.org/ and Byrd [2023a] for further detail of the Blank Center CARE^™^ Model). Notably, Camp Dream. Speak. Live. *explicitly* excludes any goals that target increased fluency and/or modification of the child’s stuttered speech. That is, speech fluency is neither a clinical target, nor an expected direct or indirect clinical outcome, and there is no reinforcement of incidental moments of reduced stuttering and/or increased fluency.

The positive cognitive, affective, and communicative outcomes of in-person administration of Camp Dream. Speak. Live. have been replicated in a series of single arm, pre-post clinical efficacy studies (Byrd et al., 2016a, 2016b, 2018, 2021). Byrd et al. (2016a, 2016b) found that children who stutter report a decrease in the adverse impact of stuttering (as measured by the *OASES*), with both children and their caregivers reporting an increase in their positive perceptions of the child’s peer relationships (as measured by the *PROMIS Pediatric Peer Relationships -Short Form 8a* [*PROMIS Peer Relationships;* DeWalt et al., 2013]). In a later study, Byrd et al. (2018) replicated these positive post-treatment gains with a cohort of school-age children who stutter (*n* = 23) and found that these gains were not dependent on pre-treatment stuttering frequency.

More recently, Byrd et al. (2021) further replicated these cognitive and affective gains with a larger sample (*n* = 37) and extended clinical efficacy outcomes to include unfamiliar listener-rated judgment of communication competencies. To do so, a speech-language pathologist who was unfamiliar to the treatment, study objectives, and participants rated participants’ impromptu oral presentations, which were video-recorded on the first and last day of treatment. Communication skills were assessed using nine core communication competencies derived from scoring rubrics from the National Communication Association (i.e., language use, organization, speech rate, intonation, volume, gesture, body position, eye contact, facial affect; Morreale et al., 2007; Spitzberg, 2007). Findings indicated significant gains in eight of the nine communication competencies, and, consistent with Byrd et al. (2018), pre-treatment stuttering frequency did not predict post-treatment gains.

Although previously conducted clinical efficacy studies of Camp Dream. Speak. Live. suggest meaningful outcomes for multiple cohorts of children and adolescents who stutter when administered in person, to date, no study has investigated the efficacy of administration in virtual format. Exploration of the outcomes following a virtual adaptation of Camp Dream. Speak. Live. holds implications for supporting children and adolescents who stutter who have minimal to no access to in-person treatment. Further, exploring the effectiveness of an approach to treatment that does not include fluency as a target would provide insight into non-ableist, whole-person approaches (Byrd, 2021; Constantino et al., 2022; Watermeyer & Kathard, 2016).

### 1.3 Purpose of this study

Although telepractice is steadily becoming a more common alternative to treatment in the field of speech-language pathology, there are relatively few studies that have explored the efficacy of providing group treatment of stuttering in a virtual format. Camp Dream. Speak. Live. is a distinct approach with demonstrated cognitive, affective, and communicative gains in children who stutter when provided in-person (Byrd et al., 2016a, 2016b, 2018, 2021). Given the forced pivot to online during the pandemic, and ASHA’s call for evidence to support the efficacy of telepractice, the present study examined whether a virtual adaptation of Camp Dream. Speak. Live. (i.e., Virtual Camp Dream. Speak. Live.) – results in similar affective, cognitive, and communicative post-intervention outcomes to those observed in in-person implementations. Specifically, the following five research questions were addressed:

**RQ1**: Does participation in Virtual Camp Dream. Speak. Live. improve self-reported communication attitude (*KiddyCAT/CAT*)?

**RQ2**: Does participation in Virtual Camp. Dream. Speak. Live. reduce the adverse impact of stuttering on quality of life (*OASES*)?

**RQ3**: Does participation in Virtual Camp Dream. Speak. Live. improve self- and caregiver-reported perceptions of peer relationships (*PROMIS Peer Relationships*)?

**RQ4**: Does participation in Virtual Camp Dream. Speak. Live. yield positive gains in communication competence as measured by an unfamiliar clinician?

**RQ5**: Does stuttering frequency prior to participation in Virtual Camp Dream. Speak. Live. predict post-treatment changes in communication competence as measured by an unfamiliar clinician?

## 2.0 Methods

### 2.1 Participants

This study was approved by the authors’ university institutional review board (IRB: 2015-05-0044) and is part of an ongoing series of registered clinical trials (clinicaltrials.gov, NCT 05908123; Byrd, 2023a) designed to examine clinical outcomes of the application of the Blank Center CARE^™^ Model (Byrd, 2023b). Written, informed caregiver permission and child assent were obtained for each participant. Sixty-one children who stutter (*n* = 1 non-binary, *n* = 14 female, *n* = 46 male) between the ages of 3 and 17 (*M* = 9.24 years, *SD* = 3.20) participated as first-time attendees of Virtual Camp Dream. Speak. Live. in one of two virtual implementations in either 2020 or 2021. Fifty-eight children participated from nine different states from the US (Delaware: n = 1; Florida: n = 1; New York: n = 1; North Carolina; n = 1; Pennsylvania: n = 2; Tennessee: n = 1; Texas: n = 49; Virgina: n = 1; Washington, n = 1). The remaining three participated virtually from Ireland in coordination with our international affiliates. All participants previously received a formal diagnosis of stuttering by a certified speech-language pathologist. Additionally, stuttering frequency was calculated via %SS based on a 200-500 syllable conversational speech sample collected the week prior to treatment (*M* = 10.07; *SD* = 8.64, range = 0 to 42). All samples were coded by a speech-language pathology clinical fellow trained in the disfluency coding protocol used in previous studies (e.g., Byrd et al., 2021, 2022). Table 1 provides demographic details of participants, including age, race, gender, ethnicity, socioeconomic status (free and reduced-price lunch: Domina et al., 2018; Nicholson et al., 2014; neighborhood affluence: Melendez et al., 2020), mono-/multilingual status, treatment history, stuttering frequency, and sampling context.

**Table 1:** Demographics of Participants in Virtual Camp Dream. Speak. Live. [Table 1 *removed per MedRxiv guidelines. Contact the corresponding author for information related to participant demographics*.]

### 2.2 Procedures

Virtual Camp Dream. Speak. Live. was administered via a secure, HIPAA-compliant university-based Zoom account in 2020 and 2021. The daily scripts for Virtual Camp Dream. Speak. Live. were adapted by the first author based on the manualized protocol for in-person administration (Byrd, 2023b), described in Byrd et al. (2016a, 2016b, 2018, 2021). The daily schedule for Virtual Camp Dream. Speak. Live. contains activities that correspond with the Blank Center CARE^™^ Model for treatment of stuttering, which targets four central components: Communication, Advocacy, Resiliency, and Education.

Several steps were taken to adapt the in-person administration to virtual administration. First, training specific to communication competence was modified to explicitly compare and/or contrast virtual communication skills to in-person communication skills. For example, participants engaged in activities where they stood at a distance from the camera while giving a presentation to replicate an in-person communication experience, versus being placed within small group breakout rooms to learn how to adjust their competencies (e.g., body positioning, gestures) relative to the communication environment.

Second, although events that incorporated speaking in different physical spaces were not possible (e.g., presenting in high-traffic public spaces, surveying unfamiliar bystanders about stuttering facts), as is traditional in the in-person implementation, measures were taken to facilitate peer-to-peer communication in a virtual space. Similar to the in-person small group assignments (Byrd et al., 2016a; 2018; 2021), participants were assigned to one mixed-age small group and one same-age small group for various activities throughout the week.

Breakout rooms were utilized across specific groups which included at least one student clinician and/or supervisor trained to specifically promote peer-to-peer communication. During large group discussions, clinicians modeled and encouraged participants to engage in peer-to-peer discussion and used the Zoom spotlighting feature to highlight individual participants or panels of participants. The ‘raise hand’ feature on Zoom was modeled by clinicians and encouraged during small and large group discussions to further facilitate interaction.

Finally, although the structure of the treatment protocol remained the same, schedule adaptations were made to optimize engagement and minimize potential screen time fatigue. Treatment days were reduced in length (6 hours/day to 3.5 hours/day), and participants were permitted to take screen breaks, if needed.

#### 2.2.1 Outcome measures

Participants completed pre-treatment and post-treatment measures collected via two Zoom sessions 3 to 7 days prior to intervention and again 3 to 7 days following intervention. Treatment outcomes included both self- and caregiver-reported measures as well as observer-rated behavioral measures. Self- and caregiver-reported measures included (a) attitudes towards communication, (b) perception of peer relationships, and (c) perception of stuttering’s impact on overall quality of life. Behavioral measures consisted of evaluations of participants’ core communication competencies by an unfamiliar clinician during impromptu presentations recorded pre- and post-treatment.

##### 2.2.1.1 Self- and caregiver-report measures

Participants between the ages of 3 and 6 years completed the *KiddyCAT* (Vanryckghem & Brutten, 2009) and participants 7 years or older completed the *CAT* (Vanryckghem & Brutten, 2009) to assess communication attitudes. Caregivers of all participants (3-17 years) completed the *PROMIS Parent Proxy* (DeWalt et al., 2013) to assess caregivers’ perception of their child’s abilities to make friends. Participants ages 8-17 completed the *PROMIS Peer Relationships* (DeWalt et al., 2013) to assess perceived ability to make friends. Participants aged 7 and older completed the *OASES* (*OASES-S*, ages 7-12; *OASES-T*, ages 13-17; Yaruss & Quesal, 2016) to assess negative impact of stuttering.

##### 2.2.1.2 Behavioral measures

To assess communication skills, participants completed impromptu presentations at pre- and post-intervention to Zoom-based audiences of three unfamiliar adults. Prior to preparation, participants were asked to introduce themselves by stating their name, where they live, their age, and to engage in an impromptu presentation from a selection of topics (see Byrd et al. 2021). Participants were provided two minutes to prepare however they preferred (e.g., writing down notes in a journal, drawing ideas, silently thinking).

All pre- and post-treatment presentations were rated by a clinician who was unfamiliar with the participants, intervention methodology, and research questions. Videos were rated across each of the nine core competencies described in Byrd et al. (2021, 2022), and the training process was identical to the methodology used in Byrd et al. (2021). In brief, the clinician was first introduced to the nine core communication competencies (i.e., language, organization, speech rate, intonation, volume, gestures, body movement, eye contact, and facial affect) and description of key features for each, while watching virtual, impromptu presentation videos unrelated to the present study with the third author, a licensed speech-language pathologist with expertise in stuttering and the present protocol. Next, the clinician and the third author each independently rated 4-6 example videos of school-age participants providing impromptu presentations in the virtual format using Qualtrics. The clinician first identified whether the behavior was present via a binary yes/no question. If not present, (e.g., if no gestures were present throughout the presentation) the communication competency received a rating of 0. If present, the clinician was presented with a 100-point visual analog scale and was instructed to move the icon along the scale to indicate the participant’s implementation of that communication skill. Higher ratings indicated more skilled performance. The clinician and third author reviewed ratings and discussed any ratings with greater than 10 points discrepancy to reach a consensus.

The clinician and the third author completed 6 sets of novel training videos and consensus discussions, resulting in inter rater reliability greater than 82% across each core competency following consensus (language: 0.98; organization: 0.99; speech rate: 0.94; intonation: 0.82; volume: 0.91; gestures: 0.97; body movement/orientation: 0.98; eye contact: 0.99; facial affect: 0.99, *p* < 0.05 for all competencies).

To evaluate the entire corpus of participant videos, the clinician used the same scaling system described above as they viewed each of the 122 presentations (61 pre-treatment, 61 post-treatment) in a randomized order and with no label of pre-/post-treatment status for any video. Each video was rated for all nine communication competencies, and no video was watched more than four times immediately prior to rating. The third author randomly assigned all participants into two groups: participants who would have their pre-treatment videos viewed before their post-treatment videos, and participants who would have their pre-treatment videos viewed after their post-treatment videos. Additionally, participants were randomized into two sequential groups such that each participant was only rated one time within the first 50% of videos viewed, and no participant was rated consecutively. The third author rated 20% of videos (24 of 122) that the clinician completed. Intra-class correlations were above 80% for eight of nine communication competencies (language: 0.93, organization: 0.97, speech rate: 0.89, speech prosody: 0.90, gestures: 0.99, body positioning/movement: 0.92, eye contact: 0.97, facial affect: 0.80, *p*-values < 0.001) and below 70% for volume (0.68, *p* < .001).

### 2.3 Statistical analyses

Paired *t*-tests were conducted to compare pre- and post-treatment ratings for the self- and caregiver-report measures (i.e.. RQ1-RQ3). To examine RQ4, nine paired t-tests were conducted to compare mean core communication competency ratings pre- and post-treatment for each of the nine competencies. Because of the multiple measurement nature of the *OASES* (RQ2) and nine communication competency ratings (RQ4), we used a Holm-Bonferroni correction for these comparisons (Holm, 1979).

For RQ1 through RQ4, all analyses used an alpha level of 0.05. Significance was confirmed using a nonparametric alternative, Wilcoxon’s signed-rank test, due to the preliminary nature and relatively modest sample size (e.g., Csibra, 2008; Farroni et al., 2005; Fasloi et al., 2004; Nettle et al., 2013; see also Byrd et al., 2021, 2022). Ties observed during nonparametric analyses were assigned mean rank values (see Moore et al., 2016). Cohen’s *d* was calculated for all significant *t* values to obtain effect sizes (Cohen, 1988).

To examine RQ5, nine linear regression models, one for each core communication competency, were conducted with pre-post difference scores as the outcome and stuttering frequency (i.e., pre-treatment %SS) as the sole predictor.

## 3.0 Results

### 3.1 Self- and caregiver-report measures

Descriptive data for self- and caregiver-report measures are summarized in Table 2. Data for self- and caregiver-reported measures of the 61 participants were included in the final analyses only if the participant (a) met the predetermined age criteria for each measure, and (b) completed each measure in full at pre- and post-treatment timepoints. All 61 participants (100%) completed the *KiddyCAT* (15 of 15 participants < 7 years old) or *CAT* (46 of 46 participants ≥ 7 years old) during pre- and post-treatment. Of the 46 participants eligible for *OASES* (≥ 7 years of age), 45 (97.8%) completed during both pre- and post-treatment. Of the 42 participants eligible for *PROMIS Peer Relationships* (≥ 8 years of age), 42 (100%) completed during both pre- and post-treatment. Of the 61 caregivers eligible for *PROMIS Parent Proxy*, 55 (90.1%) completed both pre- and post-treatment.

**Table 2:** Self- and caregiver-report Measures Pre-treatment and Post-treatment.

| Measure | Rater | N | Pre-Tx | Post-Tx | p |
| --- | --- | --- | --- | --- | --- |
| KiddyCAT | Self | 15 | 2.80 (.65) | 2.20 (.58) | .11 |
| CAT | Self | 46 | 12.84<br>(1.16) | 11.73<br>(1.06) | <.05 |
| OASES |  |  |  |  |  |
| Total Adverse Impact | Self | 45 | 2.32 (.08) | 2.12 (.07) | <.01 |
| General Information | Self | 45 | 2.67 (.07) | 2.50 (.06) | <.01 |
| Speaker's Reactions to Stuttering | Self | 45 | 2.38 (.12) | 2.09 (.09) | <.01 |
| Communication in Daily Situations | Self | 45 | 2.20 (.08) | 2.12 (.09) | .15 |
| Quality of Life | Self | 45 | 1.86 (.09) | 1.66 (.08) | <.01 |
| PROMIS Peer Relationships | Self | 42 | 47.71<br>(1.23) | 46.09<br>(1.25) | .41 |
| PROMIS Peer Relationships – Parent Proxy | Caregiver | 55 | 46.09<br>(1.30) | 48.33<br>(1.22) | <.05 |

*Note.* Pre-Tx: Pre-treatment; Post-Tx: Post-treatment; Mean and standard error of mean (in parentheses) reported for pre- and post-treatment.

**RQ1**: Does participation in Virtual Camp Dream. Speak. Live. improve self-reported communication attitude [*KiddyCAT/CAT*]?

Significant post-treatment gains were observed for the *CAT t*(45) = 2.08, *p* < 0.05, *d* = 0.31 [small-medium effect size], which reflects improved perception of communication attitudes. No significant post-treatment changes were observed for the *KiddyCAT t*(14) = 1.72, *p* = 0.11, *d* = 0.22). Figure 1 depicts the outcomes for the *CAT* and *KiddyCAT*.

**Figure 1:**
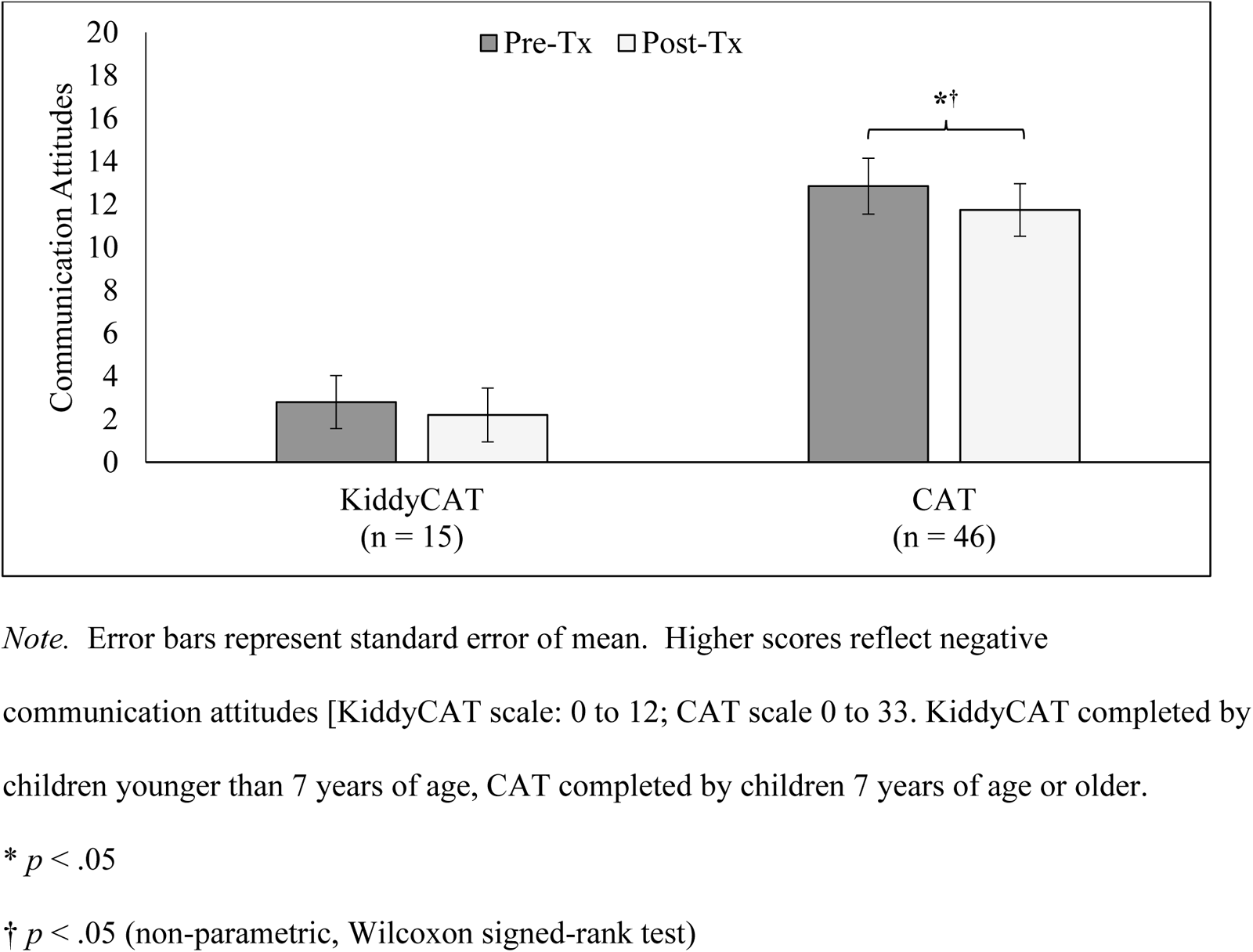
Pre- and Post-Treatment Ratings of Attitudes Towards Speaking

**RQ2**: Does participation in Virtual Camp Dream. Speak. Live. yield positive gains in impact of stuttering on quality of life [OASES]?

Significant reductions in the adverse impact of stuttering were observed post-treatment based on the *OASES* Total Impact Score *t*(44) = 5.32, *p* <.001, *d* =0.79 [large effect size], as well as three of the four subscales (General Information: *t*(44) = 3.36, *p* < .003; *d* = 0.50 [medium effect size]; Reactions to Stuttering: *t*(44) = 4.38, *p* < .001, *d* = 0.65 [small-medium effect size]; Communication in Daily Situations: *t*(44) = 1.51, *p* = 0.15, *d* = 0.23; Quality of Life: *t*(44) = 3.46, *p* < 0.01, *d* = 0.52 [medium effect size]). Figure 2 depicts the outcomes for the *OASES*.

**Figure 2:**
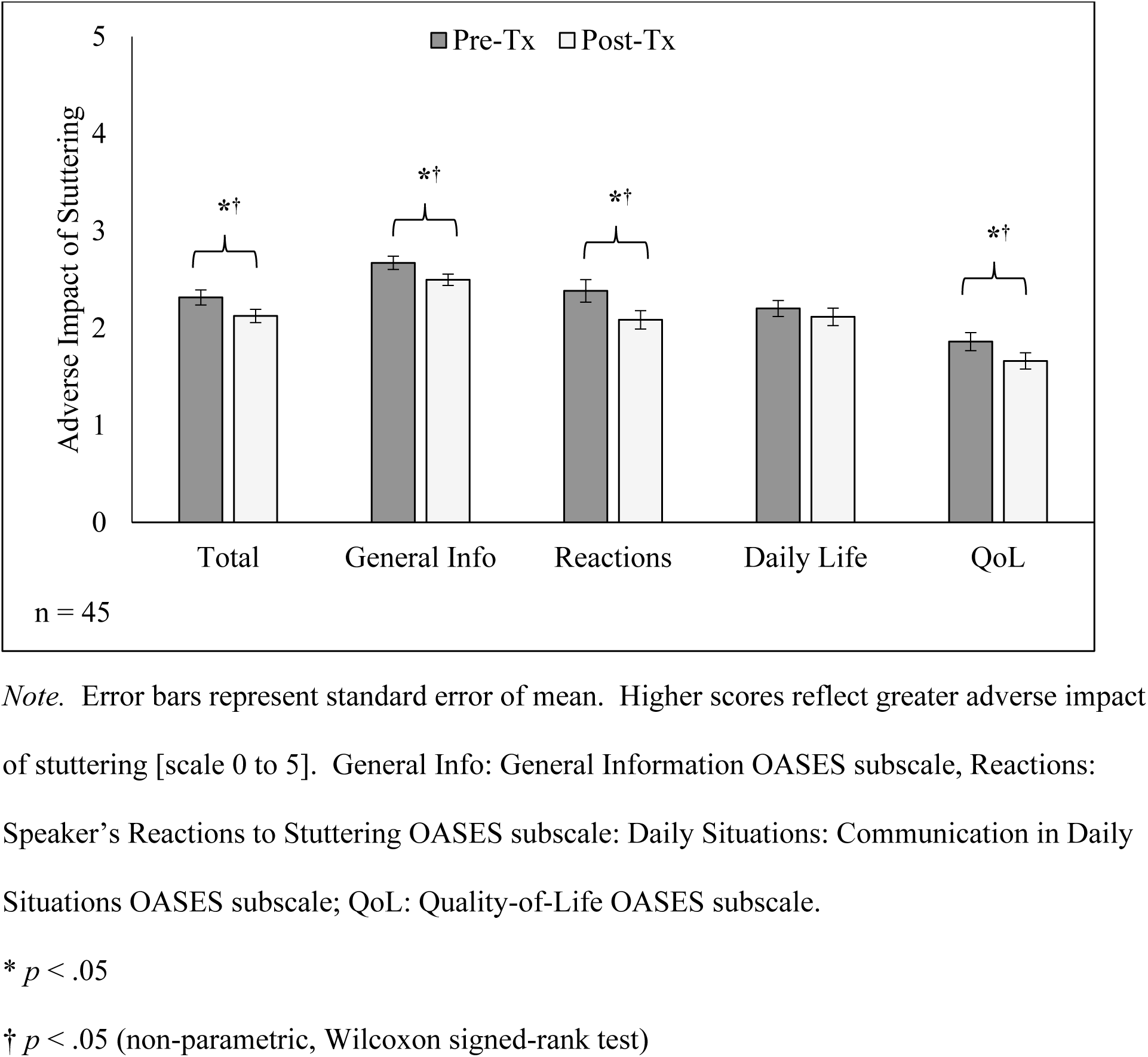
Pre- and Post-Treatment ratings of the Adverse Impact of Stuttering

**RQ3**: Does participation in Virtual Camp Dream. Speak. Live. improve self- and caregiver-reported perceptions of peer relationships and ability to establish new peer relationships [*PROMIS Peer Relationships*]?

No significant changes were observed in the *PROMIS Peer Relationships t*(41) = 0.41, *p* = 0.67. A significant increase on the *PROMIS Parent Proxy* scores *t*(54) = 2.02, *p* < 0.05, *d* = 0.27 [small effect size] was detected, which reflects improved caregiver perceptions. Figure 3 depicts outcomes for the *PROMIS Peer Relationship* scale and the *PROMIS Parent t Proxy* scale.

**Figure 3:**
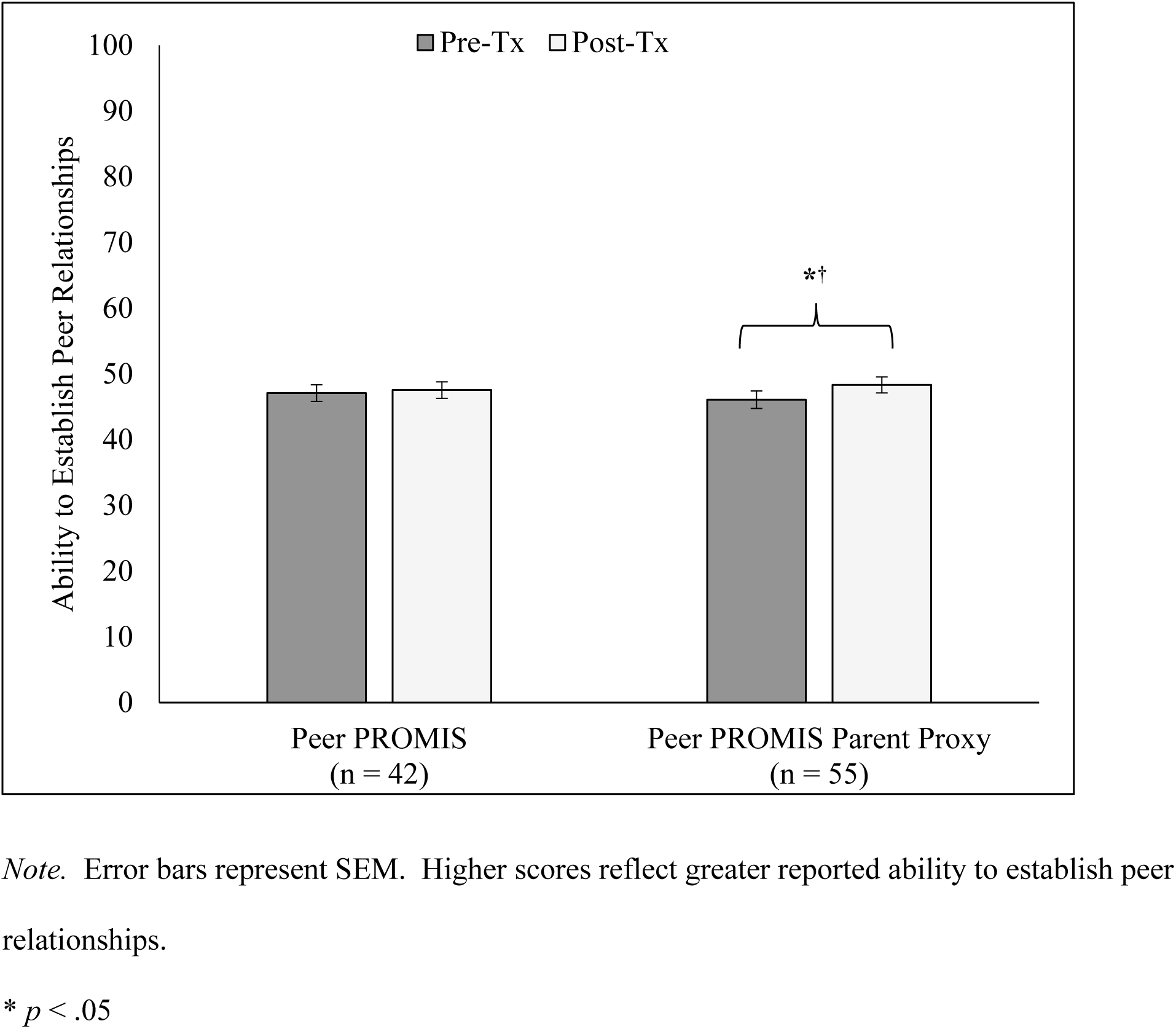
Pre- and Post-Treatment Ratings of Perceived Ability to Establish Peer Relationships

**RQ4**: Does participation in Virtual Camp Dream. Speak. Live. yield positive gains in listener-perceived communication competence?

Significant improvements were observed post-treatment in seven of the nine communication competences post-treatment, including language use (*d* = 1.41, very large effect size), language organization (*d* = 1.08, large-very large effect size), speech rate (*d* = .38, small-medium effect size), intonation (*d* = .33, small-medium effect size), volume (*d* = .45, medium effect size), eye contact (*d* = .40, small-medium effect size), and facial affect (*d* = .32, small effect size). Table 3 summarizes behavioral outcomes for each of the nine clinician-rated communication competencies.

**Table 3:**
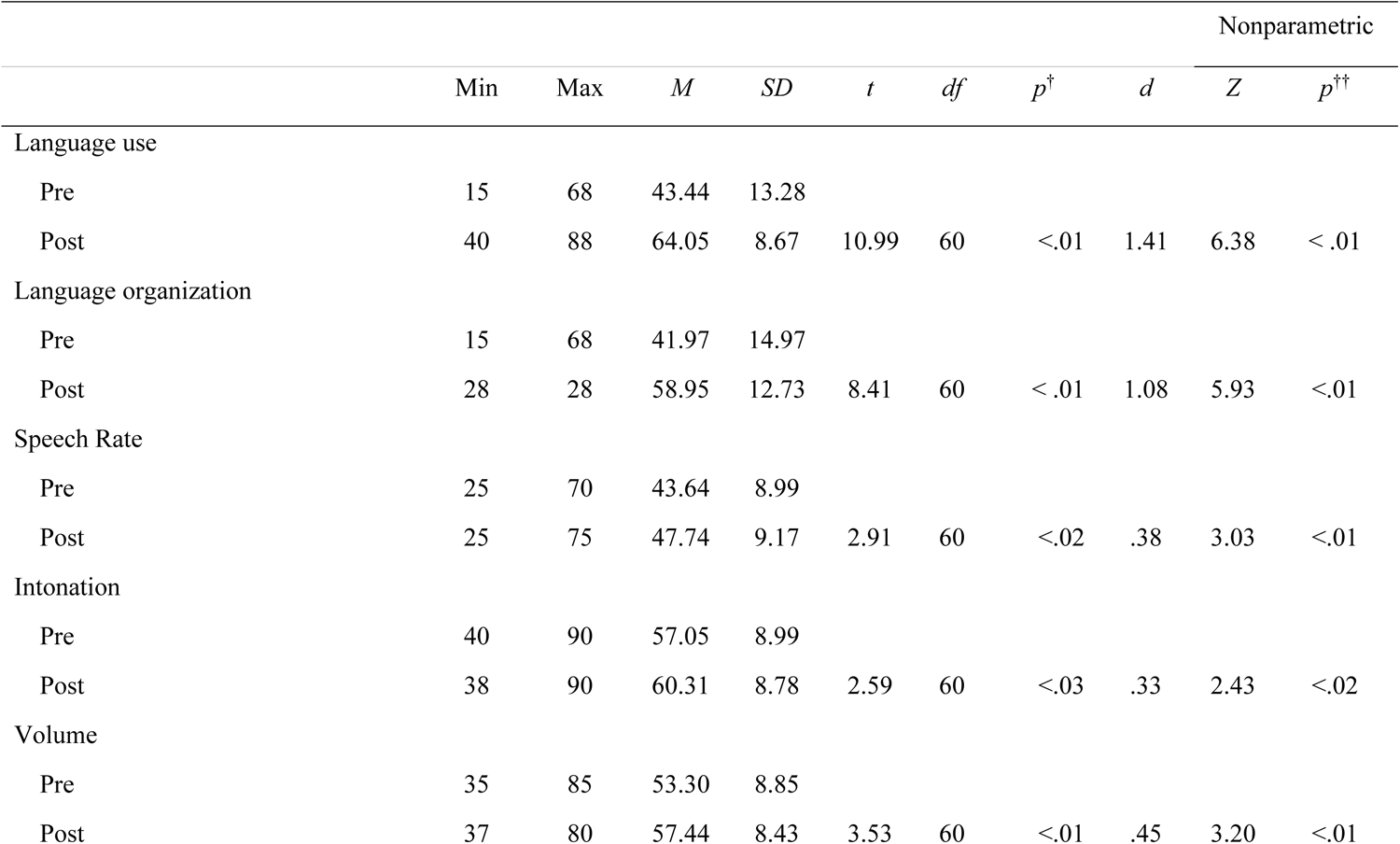
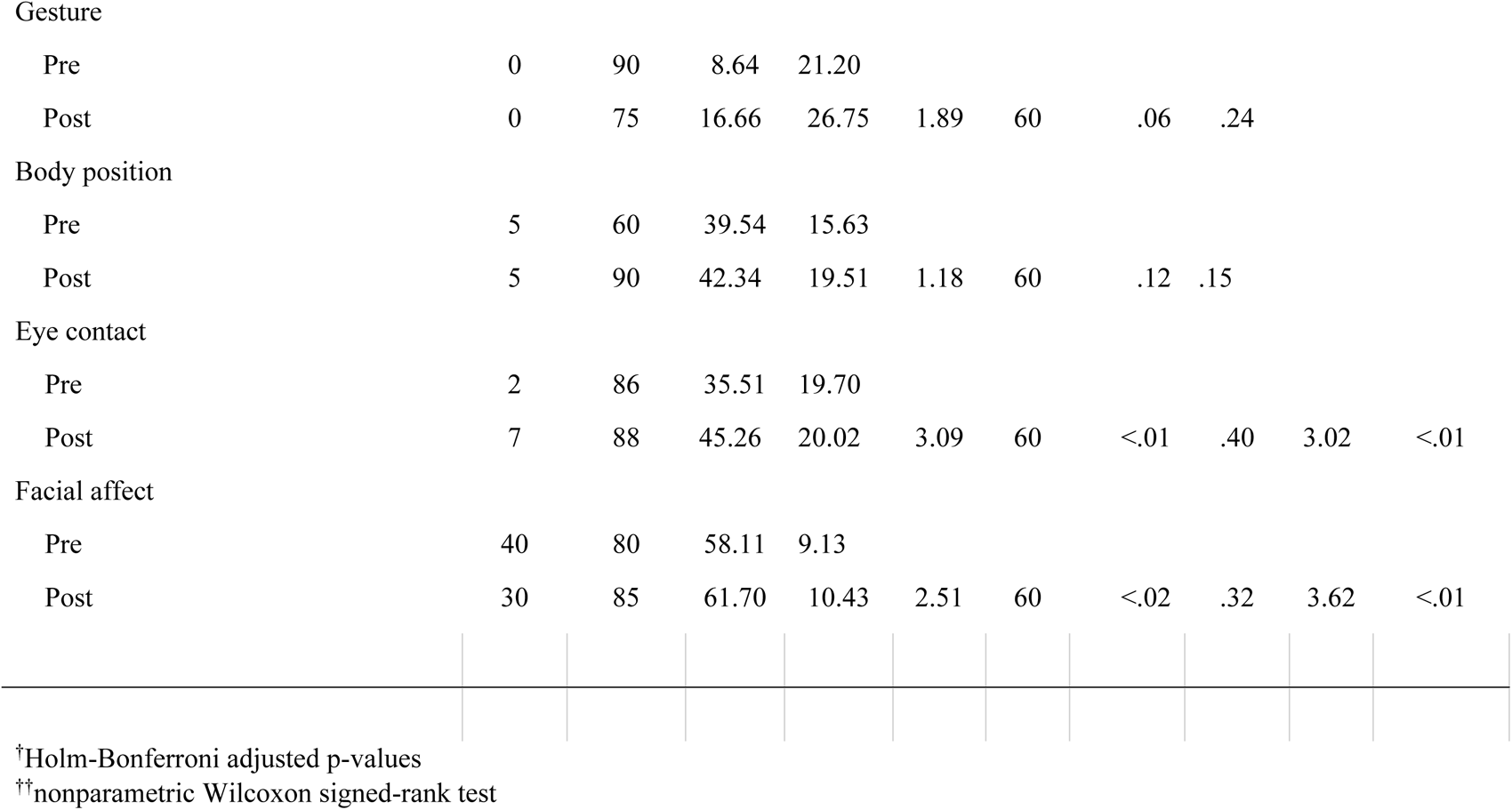
Pre- and Post-Treatment Communication Competency Scores.

|  |  |  |  |  |  |  |  |  | Nonparametric |  |
| --- | --- | --- | --- | --- | --- | --- | --- | --- | --- | --- |
|  | Min | Max | *M* | *SD* | *t* | *df* | *p*<sup>†</sup> | *d* | *Z* | *p*<sup>††</sup> |
| Language use |  |  |  |  |  |  |  |  |  |  |
| Pre | 15 | 68 | 43.44 | 13.28 |  |  |  |  |  |  |
| Post | 40 | 88 | 64.05 | 8.67 | 10.99 | 60 | <.01 | 1.41 | 6.38 | < .01 |
| Language organization |  |  |  |  |  |  |  |  |  |  |
| Pre | 15 | 68 | 41.97 | 14.97 |  |  |  |  |  |  |
| Post | 28 | 28 | 58.95 | 12.73 | 8.41 | 60 | < .01 | 1.08 | 5.93 | <.01 |
| Speech Rate |  |  |  |  |  |  |  |  |  |  |
| Pre | 25 | 70 | 43.64 | 8.99 |  |  |  |  |  |  |
| Post | 25 | 75 | 47.74 | 9.17 | 2.91 | 60 | <.02 | .38 | 3.03 | <.01 |
| Intonation |  |  |  |  |  |  |  |  |  |  |
| Pre | 40 | 90 | 57.05 | 8.99 |  |  |  |  |  |  |
| Post | 38 | 90 | 60.31 | 8.78 | 2.59 | 60 | <.03 | .33 | 2.43 | <.02 |
| Volume |  |  |  |  |  |  |  |  |  |  |
| Pre | 35 | 85 | 53.30 | 8.85 |  |  |  |  |  |  |
| Post | 37 | 80 | 57.44 | 8.43 | 3.53 | 60 | <.01 | .45 | 3.20 | <.01 |
| Gesture |  |  |  |  |  |  |  |  |  |  |
| Pre | 0 | 90 | 8.64 | 21.20 |  |  |  |  |  |  |
| Post | 0 | 75 | 16.66 | 26.75 | 1.89 | 60 | .06 | .24 |  |  |
| Body position |  |  |  |  |  |  |  |  |  |  |
| Pre | 5 | 60 | 39.54 | 15.63 |  |  |  |  |  |  |
| Post | 5 | 90 | 42.34 | 19.51 | 1.18 | 60 | .12 | .15 |  |  |
| Eye contact |  |  |  |  |  |  |  |  |  |  |
| Pre | 2 | 86 | 35.51 | 19.70 |  |  |  |  |  |  |
| Post | 7 | 88 | 45.26 | 20.02 | 3.09 | 60 | <.01 | .40 | 3.02 | <.01 |
| Facial affect |  |  |  |  |  |  |  |  |  |  |
| Pre | 40 | 80 | 58.11 | 9.13 |  |  |  |  |  |  |
| Post | 30 | 85 | 61.70 | 10.43 | 2.51 | 60 | <.02 | .32 | 3.62 | <.01 |

<sup>†</sup>Holm-Bonferroni adjusted p-values
<sup>††</sup>nonparametric Wilcoxon signed-rank test

**RQ5**: Does stuttering frequency prior to participation in Virtual Camp Dream. Speak.

Live. predict post-treatment changes in listener-perceived communication competence?

Table 4 summarizes nine separate linear regression models, one for each of the nine communication competencies conducted with pre-post communication competency difference score as the outcome variable and stuttering frequency (i.e., pre-treatment %SS; *M* = 10.07%, *Mdn* = 8.00%; range = 0.00% to 42.00%) as the single predictor. Stuttering frequency was not a significant predictor for pre-post communication competency difference scores for any of the nine competencies.

**Table 4:** Regression Models for Nine Communication Competencies with Stuttering Frequency as the Sole Predictor.

| Communication Competency | *B* | $\beta$ | SE | *df* | *t* | *p* |
| --- | --- | --- | --- | --- | --- | --- |
| Language use | -0.05 | -0.10 | 0.06 | 57 | -0.77 | 0.44 |
| Language organization | 0.01 | 0.01 | 0.11 | 57 | 0.13 | 0.89 |
| Speech rate | 0.11 | 0.13 | 0.10 | 57 | 1.05 | 0.29 |
| Intonation | 0.01 | 0.01 | 0.11 | 57 | 0.13 | 0.89 |
| Volume | -0.10 | -0.10 | 0.12 | 57 | -0.83 | 0.40 |
| Gesture | 0.01 | 0.03 | 0.03 | 57 | 0.28 | 0.77 |
| Body position | 0.02 | 0.04 | 0.06 | 57 | 0.36 | 0.71 |
| Eye contact | 0.01 | 0.03 | 0.04 | 57 | 0.27 | 0.78 |
| Facial affect | -0.07 | -0.10 | 0.10 | 57 | -0.79 | 0.43 |

## 4.0 Discussion

This study is the first to investigate Virtual Camp Dream. Speak. Live., an online adaptation of an in-person intensive intervention for children and adolescents who stutter implemented during the COVID-19 pandemic. Results replicate cognitive and affective post-intervention communication gains reported in previous studies examining in-person Camp Dream. Speak. Live. (Byrd et al., 2016a, 2016b, 2018, 2021) with regards to improved communication attitude and reduced, negative impact of stuttering among school-age and adolescent children. Results are distinct from previous studies in that improved caregiver perception of child peer relationships was observed, but improved self-perception of peer relationships was not.

### 4.1 RQs 1-3: Self-rated communication attitudes, adverse impact of stuttering, and perceived ability to establish peer relationships

Similar to in-person Camp Dream. Speak. Live., school-age and adolescents reported more positive communication attitudes post-intervention, as measured by the *CAT*. Participants younger than 7 years old did not report significantly different communication attitudes on the *KiddyCAT*. As seen in Table 2, younger participants who completed the *KiddyCAT* were limited in number (*N* = 15), and both pre-treatment scores (*M* = 2.80) and post-intervention scores (*M* = 2.20) were indicative of more positive communication attitudes that fell within the expected range for children who do not stutter (*M* = 1.79, *SD* = 1.78; Vanryckeghem & Brutten, 2009). These findings are consistent with previous investigations (Byrd et al., 2016a, 2021) and suggest both in-person and virtual implementations of Camp Dream. Speak. Live. improve communication attitudes for school-age children and adolescents who stutter and maintain positive communication attitudes for young children who stutter while also fostering growth in observable, listener-rated skills of communication competence (see Section 4.2).

Also consistent with previously published studies, school-age and adolescent participants reported post-treatment reduction of adverse impact of stuttering, as measured by the *OASES*. As discussed by Byrd et al. (2021), gains in specific subsections of the *OASES* vary across publications; however, all studies reveal significant improvement in Total Impact scores (i.e., Byrd et al., 2016a, 2018, 2021). Similar to Byrd et al. (2021), which included an age range comparable to the present study, participants reported significant gains in all subsections except Section III - Communication in Daily Situations. One interpretation of these results is that measurement of this particular subtest may not be ideal given the format of a week-long intensive program. It is possible that the time frame is too brief for participants to have the opportunity to initiate changes outside of intensive treatment, or perhaps less time to reflect on changes made in everyday communication situations within a three-week window. Long-term follow-up may be necessary to obtain a more comprehensive and naturalistic reflection of participation in Camp Dream. Speak. Live. and how it might impact day-to-day communication.

That being said, given the observed positive gains in communication, a more plausible explanation is that the timing of the treatment and assessment are not applicable to specific questions included on this particular subsection of the OASES. Virtual Camp Dream. Speak. Live. was provided during June when all participants, with the exception of the three from Ireland, were out of their traditional school environments, and thus some questions within the subtest do not apply (i.e., all of Section B of Subsection III, which refers to communication “at school”). These factors may have been heightened for the present study, as participants who completed Virtual Camp Dream. Speak. Live. did so during COVID-19 resulting in more restricted variety in day-to-day communication due to national social distancing requirements. Variability in subsections notwithstanding, replication of post-treatment changes in Total Impact scores suggests that virtual participation is comparable to in person participation in reducing the adverse impact of stuttering.

With regard to perception of ability to make friends, participation in Virtual Camp Dream. Speak. Live. yielded significant changes in caregivers’ perceptions, but no significant changes in child self-reported ability to form peer relationships. This discrepancy in caregiver and child perceptions of the change in their child’s ability to make friends may have been related to the methodological adaptations unique to the virtual format. To limit screen time fatigue, treatment was shortened from 6 hours/day (in-person) to 3.5-hours/day (virtual). Although deliberate efforts were made to facilitate peer-to-peer engagement throughout each day, this reduced overall time together may have contributed to the children not perceiving an improvement in their ability to make friends. Future iterations should explore extending the virtual time length to be more comparable to the in-person treatment. An additional consideration is that these children were navigating a pandemic, and they may have reflected differently on their ability to make friends given the restricted possibility of quality and maintenance of friendships during this particularly challenging time.

In contrast to the children’s self-report, parents of the children who participated in the present study did report a significant improvement in their perception of their child’s ability to make friends. Given the virtual format, the likelihood of caregivers being at home and having the opportunity to directly observe their child interacting virtually with peers during the treatment was high. Additionally, this observation of their children interacting with such a large cohort was likely in contrast to their daily observations of their child during the pandemic. Thus, their caregivers may have given more weight to the observation of their children interacting with other children than they would have if we were not navigating a pandemic. Alternatively, it is possible that the children did demonstrate gains, but that the children themselves did not observe these gains as their limited ability to interact with peers in the same shared physical space may have compromised their view of making friends. Having said that, the pre-treatment PROMIS data for children who stutter in the present study (*M* = 47.71; see Table 2) are not dissimilar from non-stuttering peers from a national sample population (standard score *M* = 50.00), suggesting being a child who stutters did not uniquely compromise the perception of the participants in the present study in terms of their ability to make friends. Nevertheless, the influence of Camp Dream. Speak. Live. on caregiver perceptions, as well as methods to improve both self- and caregiver-reported perceptions of peer relationships should continue to be explored in future virtual and in-person iterations.

Finally, findings indicated that significant improvements in communication attitudes (*CAT*) and reduction of adverse impact of stuttering (*OASES*) were observed for the present sample of participants, which was compiled across two separate years of Virtual Camp Dream. Speak. Live. (2020, 2021). Together, efficacy studies of Camp Dream. Speak. Live. conducted across five years (2015, [Byrd et al., 2016a]; 2017 [Byrd et al., 2018]; 2019, [Byrd et al, 2021]), and multiple cohorts of participants (*n* = 144 in total), demonstrate consistent improvements in how children and adolescents who stutter feel about their communication, and the adverse impact of stuttering on their lives. As noted in the Introduction, the existing clinical trials examining virtual treatments for stuttering have focused primarily on behavioral changes related to reduced frequency or severity of stuttered speech (e.g., Bridgman et al., 2016; Cangi & Toğram, 2020; Carey et al., 2012; Carey et al., 2014; Eslami Jahromi et al., 2022; Sicotte et al., 2003; Tomaiuoli et al., 2021). The present study extends the body of literature (Byrd et al., 2016a, 2018, 2021) and demonstrates that treatments that focus exclusively on improved cognitive, affective, and communication outcomes with no direct or indirect fluency goals can result in significant improvements in attitudes towards communication and overall quality of life.

### 4.2 RQ 4: Clinician-rated communication competence

Overall, participants demonstrated significant improvements in communication competence, as rated by an unfamiliar clinician. Of the nine communication behaviors assessed, significant improvements were observed in all but two: gestures and body movement. This study is the first to demonstrate that direct training of discrete communication competencies to children who stutter, via group treatment, is possible through virtual instruction. With respect to the non-significant improvement in gestures and body movement, it is possible that the virtual format limited the growth and/or measurement of these two specific skills. Evaluation of the use of these competencies is dependent on how much of the participant was visible within the frame. Observation of use of gestures, in particular, may be compromised, which, as summarized in Table 3, were rated lower than the eight other competencies both pre-treatment (*M* = 8.64) and post-treatment (*M* = 16.66). Making the screen size of the device, distance of child to screen, camera angle and position more uniform across participants may provide a more reliable measurement of these skills and possibly different outcomes.

### 4.3 RQ 5: Clinician-rated communication competence and stuttering frequency

Consistent with previous publications (e.g., Byrd et al., 2021, 2022), results indicate that participation in Virtual Camp Dream. Speak. Live. yielded significant improvement in communication competence regardless of pre-treatment stuttering frequency. These replicated findings across studies continue to suggest that treatment effects are observed for children with a range of stuttering frequency profiles. It is also noteworthy that, unlike most treatment studies, post-treatment stuttering frequency was excluded from analysis. The reason for this, as discussed in previous studies (Byrd et al., 2018, 2021; Coalson et al., 2024), is straightforward: the treatment did not target fluency, nor was it designed to directly or indirectly reduce stuttered speech, and previous research has demonstrated fluency and communication competency to be dissociable constructs in adolescents and adults who stutter (e.g., Byrd et al., 2024; Erickson & Block, 2013; Werle & Byrd, 2022a; Werle et al., 2021). Furthermore, the presumption that any changes in communication competence must be qualified through the lens of fluency may perpetuate a negative stereotype, particularly for those who consider stuttering as their natural manner of speech rather than a disorder that needs to be concealed or “fixed” (see Byrd, 2021; Constantino et al., 2022; Watermeyer & Kathard, 2016).

### 4.4 Broader clinical implications

Successful implementation of Virtual Camp Dream. Speak. Live. has several broader clinical implications for treatment programs for individuals who stutter. First, for speech-language pathologists who are working with children and adolescents who stutter, this study provides additional evidence for effective treatment of stuttering via telepractice. This is particularly important given that virtual learning, both in schools (e.g., Dorn et al., 2021) and within speech-language intervention, has become an ever-increasing format.

### 4.4 Limitations and future research

In the present study, inter-rater reliability above 80% was not achieved for one of the nine communication skills: vocal intensity. It is likely that the variability in measuring vocal intensity due to technological concerns (e.g., microphone levels, connectivity issues) resulted in less precise agreement compared to in-person assessment of the skill. Due to the variance in participant locations and quality of devices utilized, volume attributed to adequate vocal projection required greater consensus discussions than the other eight competencies. Vocal intensity was likely also mitigated by placement of microphones, proximity to screens, isolated ‘spotlight’ during specific events, and voluntary mute/unmute of microphones. Future studies of virtual adaptations of Camp Dream. Speak. Live., or communication competencies in virtual space, would likely benefit from standardized audio specifications and connectivity requirements, although implemented with caution, as additional technological requirements may exacerbate factors known to limit participation in telepractice (e.g., Samuels-Kalow et al., 2024; Scott Kruse et al., 2018).

## 5.0 Conclusions

Replication of positive communication outcomes for children and adolescents who stutter previously observed for in-person Camp Dream. Speak. Live. in an online format suggests that the manualized protocol is adaptable for successful virtual implementation. Modifications to programming were necessary (e.g., reduced length of screen time, breaks), and certain format-specific factors were observed (e.g., potential caregiver involvement), which may have influenced study findings. Modified measurement of communication behaviors such as gestures and body positioning may be necessary for participants who participate virtually in future clinical trials. Taken together, findings from Virtual Camp Dream. Speak. Live. replicate cognitive, affective, and communicative gains observed in in-person administrations, lending promise to adapted use, particularly in circumstances wherein in person participation is not possible.

## 6.0 Acknowledgements

This project was supported by the foundational grant support funded to the Arthur M. Blank Center for Stuttering Education and Research and endowed support provided through the Michael and Tami Lang Stuttering Institute, the Dr. Jennifer and Emanuel Bodner Developmental Stuttering Laboratory, and the Dealey Family Foundation Stuttering Clinic awarded to the first author. The authors would like to thank JoLynn Riojas for reliability coding as well as demographic data processing. We would also like to thank the child participants who stutter, as well as their families, who continue to participate in our ongoing clinical research.

## 7.0 Data Availability Statement

Datasets are available upon request.

